# The Puerto Rican Infant Metagenomic and Epidemiologic Study of Respiratory Outcomes (PRIMERO): Design and Baseline Characteristics for a Birth Cohort Study of Early-life Viral Respiratory Illnesses and Airway Dysfunction in Puerto Rican Children

**DOI:** 10.1101/2024.04.15.24305359

**Authors:** Jonathan I. Witonsky, Jennifer R. Elhawary, Celeste Eng, Sam S. Oh, Sandra Salazar, Maria G. Contreras, Vivian Medina, Elizabeth A. Secor, Priscilla Zhang, Jamie L. Everman, Ana Fairbanks-Mahnke, Elmar Pruesse, Satria P. Sajuthi, Chih-Hao Chang, Tsunami Rosado Guerrero, Keyshla Canales Fuentes, Natalie Lopez, Chris Angely Montanez-Lopez, Richeliz Alfonso Otero, Raymarie Colon Rivera, Leysha Rodriguez, Gabriela Vazquez, Donglei Hu, Scott Huntsman, Nathan D. Jackson, Yingchun Li, Andrew Morin, Natalie A. Nieves, Cydney Rios, Gonzalo Serrano, Blake J. M. Williams, Elad Ziv, Camille M. Moore, Dean Sheppard, Esteban Gonzalez Burchard, Max A. Seibold, Jose R. Rodriguez Santana

**Affiliations:** Department of Pediatrics University of California, San Francisco, San Francisco, CA; Department of Medicine University of California, San Francisco, San Francisco, CA; Centro de Neumología Pediátrica; San Juan, PR; Center for Genes, Environment & Health; National Jewish Health, Denver, CO; Department of Pediatrics and Division of Pulmonary Sciences and Critical Care Medicine, University of Colorado, Aurora, CO; Department of Medicine University of Colorado, Aurora, CO

**Author notes:** Shared first authorship (Jonathan I. Witonsky, Jennifer R. Elhawary). Shared senior authorship (Esteban Gonzalez Burchard, Max A. Seibold, Jose R. Rodriguez Santana). **Correspondence to:** Jonathan Witonsky, MD, MAS, Department of Pediatrics, University of California, San Francisco, 550 16th Street, 4th Floor, San Francisco, CA, 94143.

## Abstract

Epidemiologic studies demonstrate an association between early-life respiratory illnesses (RIs) and the development of childhood asthma. However, it remains uncertain whether these children are predisposed to both conditions or if early-life RIs induce alterations in airway function, immune responses, or other human biology that contribute to the development of asthma. Puerto Rican children experience a disproportionate burden of early-life RIs and asthma, making them an important population for investigating this complex interplay. PRIMERO, the *Puerto Rican Infant Metagenomics and Epidemiologic Study of Respiratory Outcomes*, recruited pregnant women and their newborns to investigate how the airways develop in early life among infants exposed to different viral RIs, and will thus provide a critical understanding of childhood asthma development. As the first asthma birth cohort in Puerto Rico, PRIMERO will prospectively follow 2,100 term healthy infants. Collected samples include post-term maternal peripheral blood, infant cord blood, the child’s peripheral blood at the year two visit, and the child’s nasal airway epithelium, collected using minimally invasive nasal swabs, at birth, during RIs over the first two years of life, and at annual healthy visits until age five. Herein, we describe the study’s design, population, recruitment strategy, study visits and procedures, and primary outcomes.

## INTRODUCTION

Asthma is the most common chronic condition in childhood, disproportionately affecting children from racially and ethnically minoritized backgrounds (1). Asthma is over 40% more prevalent among Black and Latino children compared to their non-Hispanic white counterparts, and Puerto Rican children face a threefold higher likelihood of having asthma and an over fivefold higher risk of asthma-related mortality (2–4). The development of asthma is influenced by complex gene-by-environment interactions that remain poorly understood (5–13). It is hypothesized that a key factor perpetuating asthma inequities among Puerto Rican children is their increased early-life exposure to environmental risk factors, particularly to asthma-genic respiratory viral pathogens, which are both ubiquitous and endemic in Puerto Rico (13–16). Irrespective of race or ethnicity, viral infections causing lower respiratory tract symptoms in the first two years of life are commonly linked to the subsequent development of recurrent wheezing and childhood asthma (17,18). In particular, infants who had bronchiolitis or wheezing induced by respiratory syncytial virus (RSV) face a 2.6-fold increased likelihood of developing asthma at six years of age (14). Furthermore, a recent prospective study found that children who remained free from RSV infection by one year of age had a 26% lower risk of developing asthma before age five than those infected with RSV by the same age (19). Similarly, children with early-life wheezing illnesses caused by human rhinoviruses (HRV) face a 9.8-fold increased risk of developing asthma at age six years (14).

Despite these findings, more studies are needed to examine the asthma risk attributable to early-life illnesses caused by RSV versus HRV, as well as the risk associated with other common respiratory viruses infecting young children. Additionally, the causative mechanisms underlying these associations still need to be determined. One plausible explanation is that infections with these respiratory viruses induce epithelial barrier dysfunction that leads to the development of asthma (20–25). Conversely, children may be born with specific airway or immune defects predisposing them to both poor responses to viral infections and an increased likelihood of developing asthma (26). If the former holds, mitigating lower respiratory tract illnesses (LRIs) could emerge as an effective strategy for preventing the onset of asthma. Supporting this notion, intervention trials focused on reducing early-life RSV infections have demonstrated efficacy in preventing asthma (27). This prospect has fueled the recent development of an RSV vaccine administered during pregnancy and an RSV antibody treatment for infants born during RSV season to block early-life acute RSV infections and inhibit asthma development (28,29).

The *Puerto Rican Infant Metagenomic and Epidemiologic Study of Respiratory Outcomes*, or PRIMERO, is a prospective observational birth cohort study of Puerto Rican children established to investigate the prevalence and role of all common respiratory viruses on the development of LRIs and asthma. Specifically, PRIMERO seeks to ascertain whether the association between early-life viral illness and asthma is rooted in deficiencies in the airway at birth or the subsequent alterations of the airway and airway mucosal immune system in response to viral infections. To achieve this aim, PRIMERO is designed to examine the molecular development and pathobiology of the airway at birth and throughout early childhood, all while performing surveillance for early-life respiratory illnesses, with viral and molecular response characterization of the detected illnesses. In contrast to prior birth cohort studies, our study utilizes cutting-edge multiplexed molecular assays, which allow us to identify illnesses caused by both RSV and HRV, in addition to metapneumovirus, coronaviruses, parainfluenza, influenza, and other species that have been less studied in this context. Furthermore, leveraging Puerto Rican’s rich genetic admixture, PRIMERO children will be genetically characterized to determine if the effects of these viruses on LRI and asthma risk are dependent on genetic variation. Perhaps most innovative, the PRIMERO study will characterize molecular airway dysfunction repeatedly at birth, yearly throughout childhood, and during respiratory illnesses, through transcriptomic analyses of nasal airway mucosa, collected by swabbing the inferior nasal turbinate. Importantly, our prior work in young children has shown this minimally invasive technique collects both epithelia and airway immune cells, and that the cell types and expression profiles of these upper airway samples mirror those in the lower (bronchial) airways in health and asthma disease (30). Leveraging this rich dataset, our goals are fourfold: [1] identify the genetic and viral determinants of early-life LRIs, [2] elucidate the influence of LRIs caused by different viral species on the development of asthma, [3] delineate the trajectories of airway molecular and cellular development from birth through childhood in both children who do and do not develop asthma, and [4] ascertain whether the airway developmental trajectories leading to asthma are predetermined at birth or influenced by early-life viral LRIs. Beyond these goals, PRIMERO’s unique and comprehensive characterization of respiratory illnesses and airway pathobiology during childhood offers unprecedented opportunities for further investigation into the impact of prenatal and postnatal social, environmental, and genetic determinants of childhood health and disease.

PRIMERO, a National Heart, Lung, and Blood Institute (NHLBI) funded longitudinal birth cohort consisting of 2,100 term healthy infants, is the result of a collaborative effort among research groups from the University of California, San Francisco (UCSF), National Jewish Health (NJH) in Denver, CO, and Centro de Neumología Pediátrica (CNP) in Caguas, Puerto Rico. The study meticulously tracks this cohort from birth through early childhood, incorporating RI surveillance during the initial two years of life and annual airway sampling during healthy child study visits. Herein, we provide a comprehensive overview of the objectives, study design, procedures, specimen collections, and recruitment results.

## METHODS

### Primary Hypotheses

RIs caused by some viral species will independently increase the risk of acute lower respiratory tract symptoms and asthma, while the effect of other species on these outcomes will be dependent on host factors, including genetics. Early-life viral LRIs will directly contribute to the risk of childhood asthma by modulating the early-life development of the airway epithelium and immune system.

### Primary Objectives

To address our hypotheses, PRIMERO pursues the following seven study objectives:

1. Identify the genetic and viral determinants of early-life upper RIs (URIs) and LRIs.
2. Determine the association between early-life RIs attributed to various viral species and both the modified asthma predictive index (mAPI) and asthma disease in childhood.
3. Determine whether the risk of asthma conferred by different viral RIs is dependent on host factors, including genetics.
4. Identify distinct early-life airway gene expression patterns and cellular developmental trajectories that contribute to asthma in childhood.
5. Determine whether early-life viral LRIs alter airway developmental trajectories.
6. Determine whether the molecular and cellular profiles of the airway at birth are associated with the occurrence of early-life viral LRIs and/or the development of asthma in childhood.
7. Determine whether the gene expression responses to viral LRIs are associated with the subsequent development of asthma.

### Study Overview

To address these objectives, we designed PRIMERO as a prospective observational birth cohort study, enrolling newborns and their mothers from March 2020 to June 2023. Surveillance for RIs during the child’s first two years involves weekly short message service (SMS) texts and email messages, complemented by in-person visits and biospecimen collections specifically designated for RIs. In addition, newborn and annual follow-up visits for the child’s first five years of life are used to assess respiratory health, environmental and social exposures, measure lung function, and to collect biospecimens. The minimum duration of participant follow-up will be two years with the aim to follow participants for a maximum of 10 years pending ongoing support of the study.

### Established Community Engagement

PRIMERO builds upon our extensive community engagement with the Puerto Rican community, which commenced in 1998 with the initiation of the Genetics of Asthma in Latino Americans (GALA) Study (31). This endeavor was designed to explore the heightened rates of asthma prevalence and morbidity among Puerto Rican children. Expanding on this groundwork, our research progressed with the Genes-environments & Admixture in Latino Americans (GALA II study) spanning from 2008 to 2014 (32). This case-control study focused on asthma in Latino children, including Puerto Rican participants in our recruitment, epidemiological, and genetic analyses. Our efforts in GALA and GALA II resulted in over an 85% recruitment rate and a follow-up contact rate exceeding 90% among Puerto Rican study participants, with GALA II providing much of the preliminary data for PRIMERO.

### Study Population and Recruitment

PRIMERO recruited pregnant women who were receiving obstetric/gynecologic (OB/GYN) services and giving birth at Hospital Interamericano de Medicina Avanzada-San Pablo (HIMA). HIMA is located in Caguas, Puerto Rico, which neighbors San Juan, Puerto Rico’s capital and most populous municipality (**Figure 1**). Most babies born at HIMA (>95%) come from families residing in the San Juan-Bayamón-Caguas metropolitan area, home to 63% of Puerto Rico’s population. Demographically, the San Juan-Bayamón-Caguas metropolitan area is similar to the entire island in terms of age, marital status, birth rate, and educational attainment (33).

**Figure 1.**
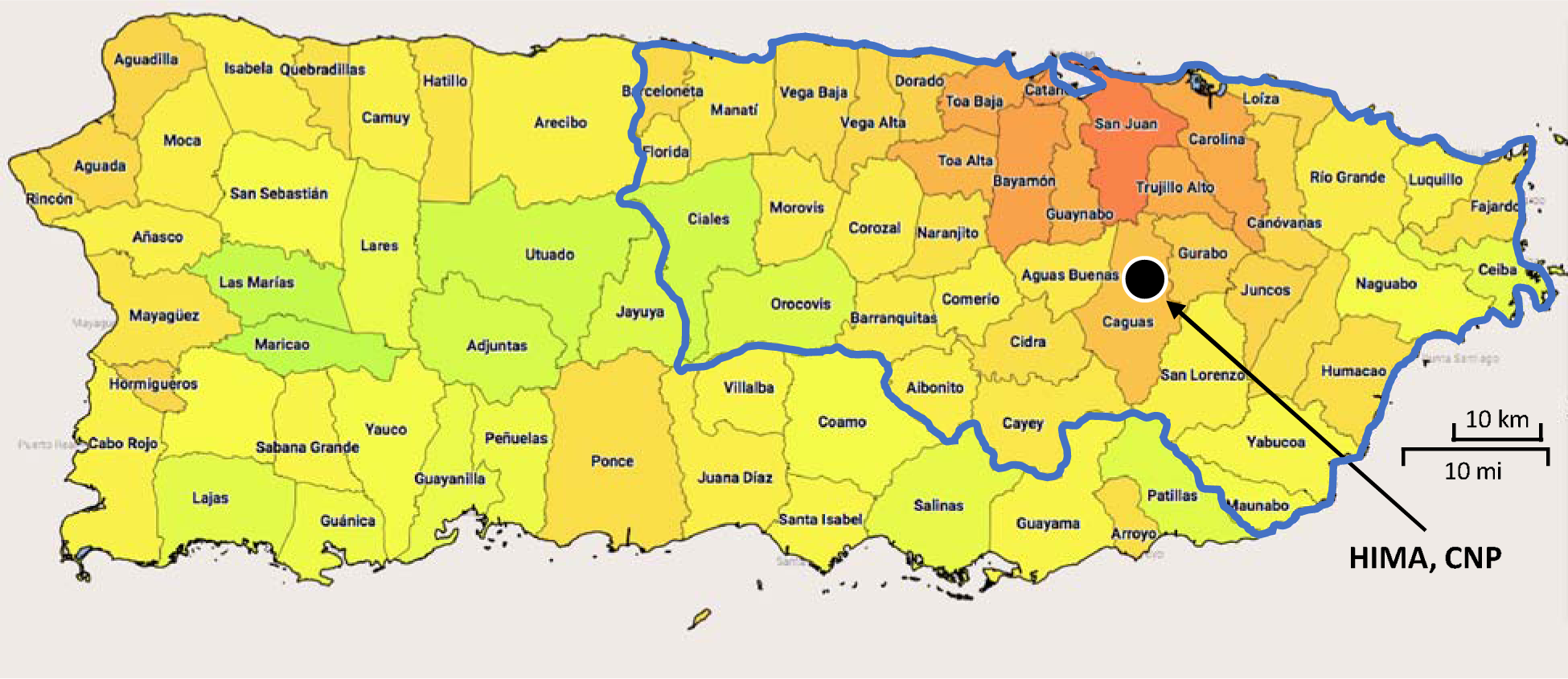
Municipalities and population density of Puerto Rico Redder areas represent municipalities with higher population densities. Greener areas represent lower population densities. The San Juan-Baymón-Carolina metropolitan area, bordered in blue, represents HIMA’s catchment area.

Healthcare providers from HIMA’s OB/GYN services were introduced to PRIMERO via presentations and information sessions. Medical staff were invited to join recruitment efforts, and collaborating offices received PRIMERO brochures for distribution during routine prenatal visits. Women receiving information packets had their details recorded in the custom electronic database, PRIMERO-DB (**Supplemental Text 1**). Recruitment occurred in two stages, allowing participants time to consider involvement. Initially, phone calls assessed interest, followed by consent and eligibility determination. PRIMERO recruiters at the HIMA labor and delivery ward used PRIMERO-DB to cross-reference consented participants. The second consent stage occurred after the child was born pre-discharge when mothers were allowed to reaffirm their participation. Protocols and documents were approved by the UCSF Institutional Review Board (IRB) and a National Institutes of Health-appointed Observational Study Monitoring Board (OSMB), ensuring data safety, study progress, and confidentiality protocols.

### Enrollment, Study Visits, and Procedures

Study visits and procedures are detailed in **Table 1**. Briefly, study participation consists of an enrollment visit, annual visits for the first five years of life, and recurring RI visits during the first two years of life.

**Table 1.**
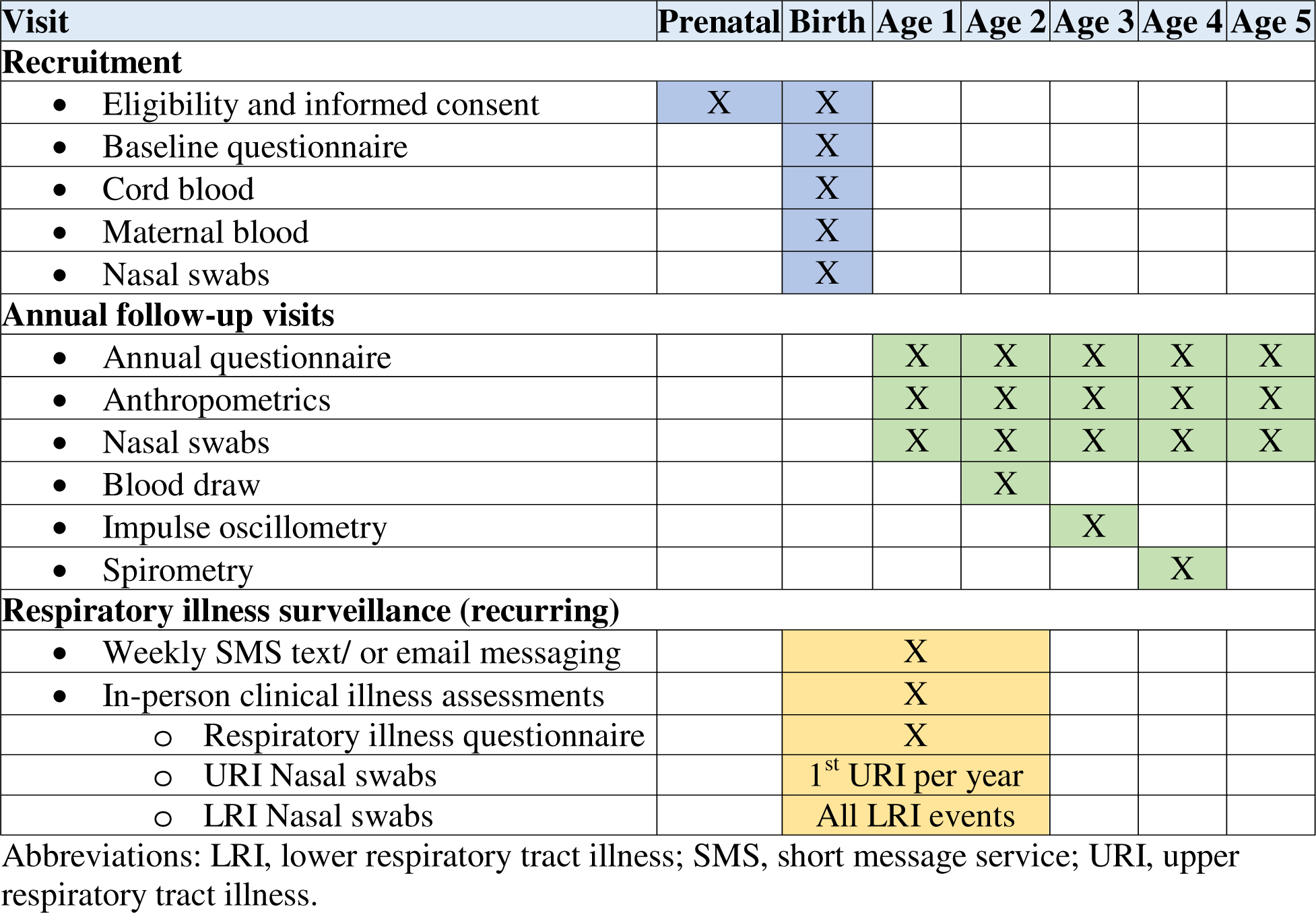
Schedule of study procedures and visits.

#### Enrollment

Before birth, expectant mothers in the HIMA OB/GYN clinic expressing interest in the study were assessed for eligibility and consented. Inclusion and exclusion criteria are provided in **Table 2**. At birth, the newborns of consented mothers were assessed for eligibility, and cord blood was collected. In the recovery ward of HIMA hospital, mothers of eligible infants were approached with a reconsent. Reconsented mothers were then administered a baseline questionnaire by trained staff who also collected peripheral blood from the mother and nasal swabs of the inferior turbinates of the newborn. The cord blood collected from infants whose mothers did not reconsent and those infants deemed not eligible was discarded, and infants were not enrolled into the study. Blood and nasal samples were processed at CNP, the core facility for our recruitment and follow-up procedures, located in the building adjacent to HIMA. After processing, samples were shipped to each of the laboratory cores: blood to the UCSF Pediatric Asthma Biobank and nasal swabs to the NJH Nasal Airway Biobank (**Supplemental Text 2**).

**Table 2.** PRIMERO Enrollment Inclusion and Exclusion.

|  | **Inclusion** | **Exclusion** |
| --- | --- | --- |
| Mother | Residential address in Puerto Rico |  |
|  | At least 18 years of age |  |
|  | Have SMS text-enabled phone or working email address |  |
|  | Able to read and understand English or Spanish |  |
|  | Delivery at HIMA |  |
| Newborn | Lives with biological mother | Sibling of a child already enrolled in PRIMERO |
|  | At least 37<sup>a</sup> weeks gestational age | Multiple birth<sup>c</sup> |
|  | Birthweight of at least 2,500 g<sup>b</sup> | Requires mechanical ventilation |
|  | Afebrile (<38°C) | Immune compromised |
|  | No clinical signs of respiratory infection | Congenital<sup>d</sup> or neuromuscular<sup>e</sup> disorders |

Abbreviations: SMS, short message service; HIMA, Hospital Interamericano de Medicina Avanzada-San Pablo; g, grams; C, Celsius
<sup>a</sup> Newborns at least 36 weeks gestational age and <sup>b</sup> weighing within 10% of the 2,500 g cutoff (i.e., weighing at least 2,250 g) at birth are evaluated for inclusion on a case-by-case basis by an obstetrician/pediatrician who determined by physical examination that the child was otherwise healthy.
<sup>c</sup> Only one child of a multiple birth is allowed to enroll.
<sup>d</sup> Examples include pulmonary abnormalities and Down syndrome.
<sup>e</sup> Impaired ability to clear airway secretions.

#### Annual Visits

Initially, annual visits and questionnaires were aligned with the child’s birthday. However, due to the COVID-19 pandemic, the window was extended to 8 months post-birthday to accommodate stay-at-home mandates and vaccination rollouts. For safety and efficiency, questionnaires are conducted separately by trained PRIMERO staff via phone, utilizing the secure UCSF Research Electronic Data Capture (REDCap) software (34,35). Questionnaires administered at newborn and annual follow-up visits during the child’s initial five years gather comprehensive data on environmental factors, demographic variables, social aspects, and clinical risk factors associated with recurrent wheezing and asthma. Biospecimens are collected annually during healthy periods, and the mAPI is assessed at 2 years. Annual visits include recording anthropometrics, an eczema assessment, and collection of two nasal swabs. In the second year, peripheral blood is processed for allergen sensitization. In the third year, impulse oscillometry is conducted with the plan for spirometry in subsequent years to evaluate lung function. Annual nasal swabs are sent to the NJH Nasal Airway Biobank, and year two blood samples go to the UCSF Pediatric Asthma Biobank after processing at CNP.

#### Respiratory Illness Surveillance

After discharge from the hospital following birth, participants enter the RI surveillance phase of the study (**Figure 2**), which lasts for the first two years of life. Mothers of participating infants receive a weekly automated SMS text or email that provides a hyperlink to indicate whether the child has signs of RI (**Supplemental Figure 1**). Text and email messages that are unanswered within 24 hours or have an affirmative response are compiled into a weekly call list, followed by a phone call from project staff. In addition to active surveillance, mothers can call PRIMERO staff or visit the clinic for a phone-based or face-to-face assessment. Illness assessments, visits, follow up, and severity determination are further described in **Supplemental Text 3**.

**Figure 2.**
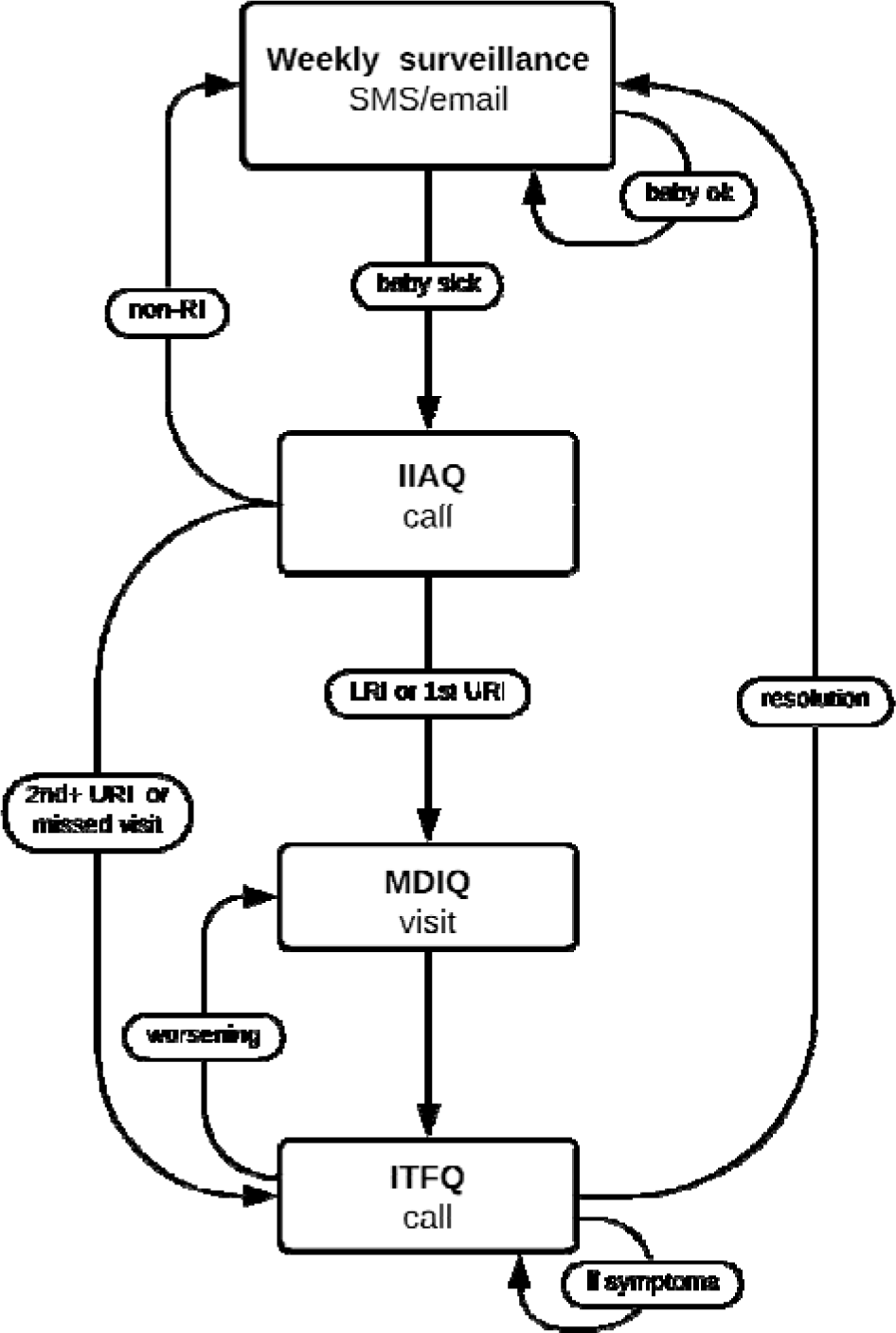
Illness Surveillance Flow Diagram Abbreviations: IIAQ, Initial Illness Assessment Questionnaire; ITFQ, Illness Tracking and Follow-up Questionnaire; LRI, lower respiratory tract illnesses; MDIQ, Medical Doctor Illness Questionnaire; non-RI, non-respiratory tract illness; SMS, short message service; URI, upper respiratory tract illness. Participants move through statuses assigned at each node to track the illness from onset until resolution.

### Biospecimen Collection

Cord and maternal blood are collected at enrollment to define genome-wide genetic variation in mothers and children. Nasal swabs are collected from newborns at enrollment, at each annual visit, and during in-person RI visits. Illness swabs are tested for SARS-CoV-2 and enterovirus D68 using PCR assays and for 22 other common respiratory pathogens using the NxTAG Respiratory Pathogen Panel. Nasal swabs are also used to evaluate longitudinal airway gene expression in health and illness using whole transcriptome RNA-sequencing. In addition, blood is collected at the year 2 visit to obtain complete blood counts with differentials and assess allergen sensitization via specific IgE testing to perennial aeroallergens and foods.

### Respiratory Illness Outcomes

RI outcomes of the study include the following:

1. Identification of early-life RIs.
2. Analysis of RI-associated airway gene expression.
3. Determination of respiratory virus infection at a species level.
4. The modified asthma predictive index (mAPI).

Secondary RI outcomes and the secondary research questions we aim to address with these outcomes are described in **Supplemental Text 4.**

#### Identification of Children with Early-life LRIs

Participants will be classified at the end of their 2-year surveillance period based on the severity of LRI(s) experienced: did not experience an LRI; experienced at least one mild/moderate LRI, but not a severe LRI; or experienced at least one severe LRI. In most cases, LRIs in the first two years of life are determined by a physician assessment. However, due to missed visits for some participants, a subset of LRIs is only defined by the symptoms reported during the screening and tracking of an illness event.

#### Analysis of Respiratory Illness-Associated Airway Gene Expression

We will use RNA sequencing to measure whole transcriptome nasal airway epithelium gene expression patterns at birth, during defined URI and LRI events, and from annual follow-up visits from ages one through five.

#### Determination of Respiratory Virus Infection at a Species Level

Viral species-specific data generated from nasal airway swab RNA and metagenomic analysis of RNA-sequence data will allow us to determine if a participant has been infected with a respiratory virus and determine if viral species are associated with illness severity outcomes.

#### mAPI

Children undergo clinical assessment for asthma risk at two years of age via the mAPI, which is an updated iteration of the asthma predictive index (API) and provides an assessment for future asthma risk (36). The mAPI has greater sensitivity than the API and has been included in the National Asthma Education and Prevention Program (NAEPP) Expert Panel Report 3 (EPR-3) guidelines as a criterion for the institution of long-term therapy to decrease asthma morbidity and exacerbations (37).

### Risk Mitigation Plan

Study enrollment commenced in March 2020, coinciding with the onset of the COVID-19 pandemic. After considering the risks of pandemic viral exposures to staff and participants, PRIMERO leadership, in conjunction with HIMA, NHLBI, and the OSMB, decided to proceed with the study, operating under a risk mitigation plan (**Supplemental Text 5**).

### Participant Engagement

To enhance retention and engagement, mothers are initially reimbursed for parking costs at HIMA during recruitment, and they receive a thank-you gift at the conclusion of the baseline and each annual follow-up visit. Beyond study visits, participants are regularly contacted through various channels, including SMS texts, email, and the study website (**Supplemental Figure 2**). The study website and newsletters serve to keep participants informed about the study’s progress and provide prompts for updating contact information. Weekly contacts during the first two years help identify RIs, and personalized SMS texts with a link to the study website are sent on the child participants’ birthdays. Additionally, participants receive annual holiday greetings via email.

## RESULTS

### Recruitment

Enrollment began in late February 2020 with the first eligible infant born in early March, coinciding with the first reported cases of COVID-19 in Puerto Rico, at which point the governor of Puerto Rico ordered an island-wide curfew and closure of non-essential businesses in mid-March 2020 (38). As additional pandemic control measures ensued, PRIMERO remained one of few studies that was successfully able to actively recruit through the height of the pandemic and continued enrolling until June 2023. Over the 40-month recruitment period, HIMA delivered 3,543 babies. Mothers who delivered over the weekends or received prenatal care outside of HIMA were not considered for recruitment. We approached 2,723 of those mothers with welcome packets and screened them for potential enrollment. Of those, 318 mothers declined participation, 183 mothers were ineligible, 12 did not reaffirm their consent, one birth was missed, and 109 infants were not eligible, culminating in a total of 2,100 mother-child dyads being enrolled in the study (**Figure 3**) representing 59% of all of HIMA births.

**Figure 3.**
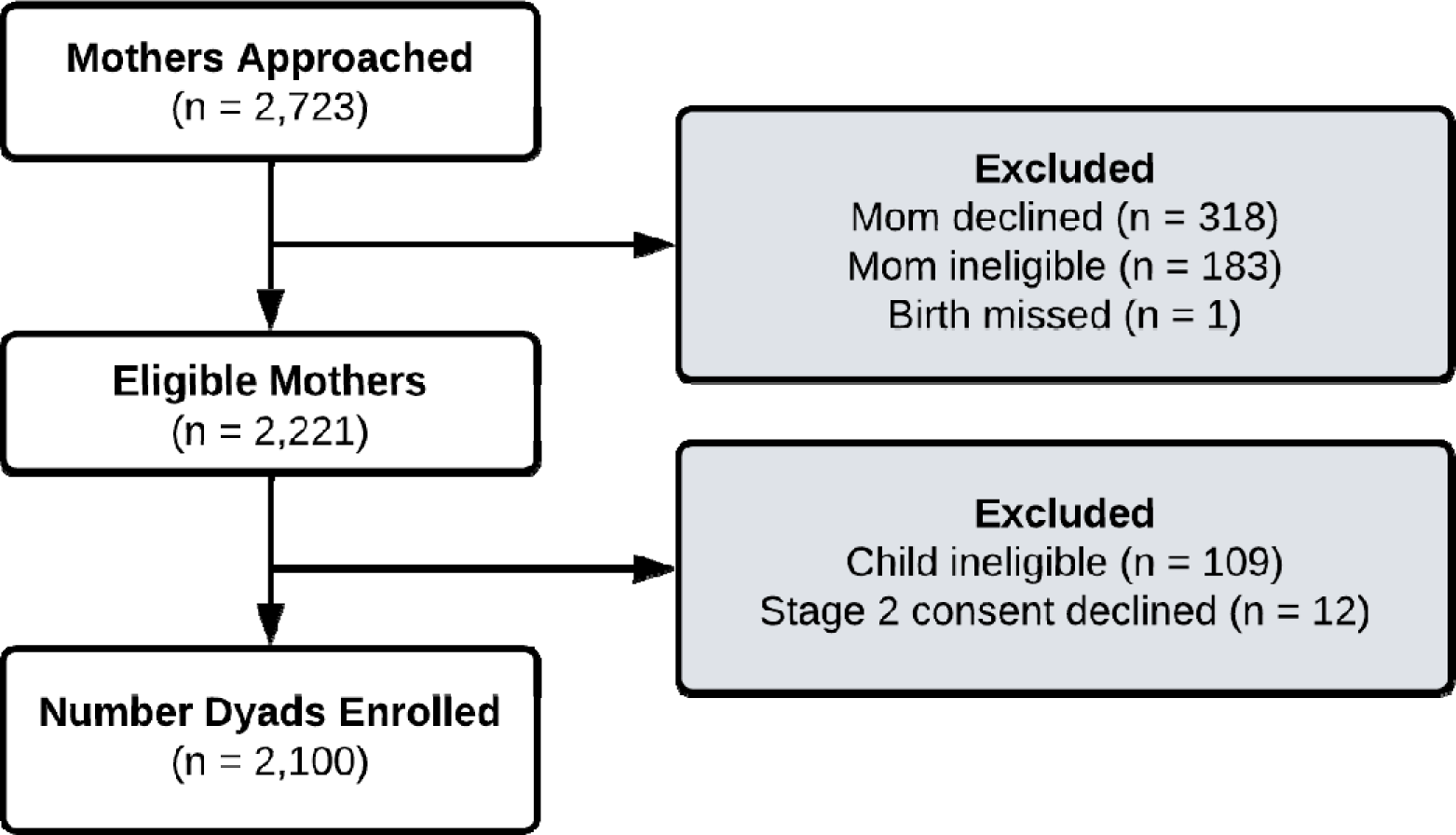
Flowchart of PRIMERO enrollment spanning February 2020 to June 2023.

### Descriptive Characteristics of Participants

Characteristics of PRIMERO mothers and children (**Table 3**) were derived from a baseline questionnaire and infant eligibility form. The median age of mothers was 26.6 (IQR, 23.1–30.7) years. Cesarean delivery was the predominant (72.6%) delivery method, with most (53.9%) unplanned. The race of each child was derived from the parents’ reported races, with 39.0% White if both parents identified as White, 22.0% Black/African if both parents identified as Black/African, and 39.0% Other/Mixed if one parent identified as White and the other as Black/African, encompassing all possible combinations (39).

**Table 3.** Characteristics of Enrolled Mothers and Children in PRIMERO, Puerto Rico, as of February 3, 2024.

| Characteristic | No. of Participants | % | Median (IQR) |
| --- | --- | --- | --- |
| Maternal age, years |  |  | 26.6 (23.1 – 30.7) |
| Cesarean deliveries | 1,524 | 72.6 |  |
| Unplanned Cesarean | 821 | 39.1 |  |
| Gestational age, weeks |  |  | 38.7 (38.0 – 39.4) |
| Birth weight, grams |  |  | 3,202 (2,915 – 3,466) |
| Child's sex, male | 1,047 | 49.8 |  |
| Child's race<sup>a</sup> |  |  |  |
| Black/African | 461 | 22.0 |  |
| Other/Mixed | 818 | 39.0 |  |
| White | 820 | 39.0 |  |
| Not specified | 1 | 0.0 |  |
| Perinatal antibiotics exposure<sup>b</sup> | 433 | 20.7 |  |
| Maternal prenatal smoking<sup>c</sup> | 22 | 1.1 |  |
| Maternal education status, low<sup>d</sup> | 739 | 35.2 |  |
| Paternal education status, low<sup>d</sup> | 946 | 45.1 |  |

Abbreviations: IQR, interquartile range
<sup>a</sup> Child's race as a derivation of parents' reported races: "White" if both parents identified as White, "Black/African" if both parents identified as Black/African, and "Other/Mixed" if one parent identified as White and the other as Black/African.
<sup>b</sup> Perinatal antibiotic exposure if neonate was administered antibiotics in 3<sup>rd</sup> trimester or neonate was administered antibiotics soon after birth.
<sup>c</sup> Maternal prenatal smoking if mother reported smoking during pregnancy or reported time from quitting is less than child's gestation age.
<sup>d</sup> Low educational status if reported highest level of education as "Less than high school education" or "High school education or GED."

### Annual Visits and Biospecimen Collection

Among children eligible for annual visits, 57% of expected visits occurred and 62% of expected questionnaires were completed as of February 3, 2024. Nasal swabs were successfully collected from 99% of these children, and blood samples were obtained from 97%. Among children three years and older who attended their third annual visit, 54% have completed impulse oscillometry.

### Participant Retention

As of February 3, 2024, 1,940 study participants remain actively engaged, while 136 are on standby status whereby the participant requested reduced contact frequency, relocated from Puerto Rico, or informed us of an inability to commit to future follow-up. Additionally, 23 participants withdrew from the study and one child died due to reasons unrelated to the study.

### Adverse Events

Related adverse events are defined as any unfavorable medical occurrence attributed to a PRIMERO procedure, such as blood draw, nasal swab, or pulmonary function testing. A singular adverse event was recorded, involving a syncopal episode during a child’s blood draw.

## DISCUSSION

The *Puerto Rican Infant Metagenomic and Epidemiologic Study of Respiratory Outcomes* (PRIMERO) was designed to unravel the early life social, environmental, and genetic contributors to severe respiratory wheezing and subsequent asthma. A primary goal is to characterize the relationship between viral RIs in early life and the subsequent development of childhood asthma, focusing on the pivotal role of airway epithelial dysfunction. PRIMERO sets out to determine whether this dysfunction manifests at birth or emerges as a consequence of viral RIs during infancy and early childhood. Our prospective birth cohort study is poised to yield groundbreaking insights into the genetic and viral risk factors associated with severe RIs. Furthermore, it aims to pinpoint distinct airway cellular developmental trajectories observable in children at high risk for asthma, both at birth, following RIs, and during periods free of respiratory symptoms preceding the onset of asthma.

"*Primero*" means "first" in Spanish, signifying PRIMERO as the inaugural asthma birth cohort study in Puerto Rico. The imperative for comprehensive research across diverse genomes cannot be overstated as it is fundamental to advancing science and medicine (40,41). Despite significant attention and resources directed toward genetic research, globally diverse populations remain notably underrepresented (42). Here, PRIMERO is helping to bridge this gap towards equal representation of study populations in biomedical science. The emphasis of PRIMERO on the population of Puerto Rico is emblematic of the Strategic Vision of NHLBI, which underscores the exploration of factors contributing to health disparities among diverse populations (43). Additionally, recruitment and follow-up activities are exclusively conducted within Puerto Rico, in close partnership with local investigators, clinicians, and other Puerto Rican professionals, enriching the biomedical workforce with diversity and expertise.

A fundamental strength of PRIMERO lies in its innovative approach, capitalizing on the minimally invasive nature of nasal airway swabs to procure critical respiratory samples at multiple times: at birth, during early-life RIs, and annually for the first five years of life (30,44,45). Our study utilizes cutting-edge multiplexed molecular assays, enabling the identification of a spectrum of RI-causing viruses, including RSV, HRV, metapneumovirus, coronaviruses, parainfluenza, influenza, and other less studied species. Collected specimens from PRIMERO participants undergo genetic characterization, allowing us to determine the role of genetic variation in the effects of these viruses on LRI and asthma. Most notably, through transcriptomic analyses of nasal airway mucosa, the PRIMERO study introduces a groundbreaking approach by longitudinally characterizing molecular airway dysfunction at birth, annually throughout childhood, and during RIs in the first two years of life. Extensive research substantiates the effectiveness and reliability of nasal swabbing, underscoring critical findings: it reveals that the upper and lower airways share similar cell types and expression patterns, and upper airway insults can elicit lower airway symptoms and immune responses (30,46–49). The inflammatory endotypes of asthma observed in both the nasal and bronchial airways exhibit a high degree of correlation, emphasizing the importance of the nasal airway epithelium as a primary site for infection with asthma-associated respiratory viruses (30). Furthermore, we previously demonstrated the versatility of RNA-sequence data generated from nasal airway epithelial swabs, serving a dual purpose: elucidating host transcriptional profiles and screening samples for pathogenic respiratory viruses through metagenomic analysis (50). This single-swab RNA-sequence approach offers a robust and tightly controlled assessment of the host response to viral infection and stands as a central analysis method within our study.

As with any complex study, PRIMERO has its unique challenges. One challenge was the low birth rate in Puerto Rico, which, at 0.90 births per woman, is approximately half that in the mainland U.S. (51). Despite the relatively small source population of potential participants, PRIMERO recruitment has been overwhelmingly successful. Another challenge was the lack of available software to manage the recruitment, surveillance, and follow-up of 2,100 mother-child dyads and to interact with their electronic medical record (EMR) as they progress through the study. A key innovation was the creation of PRIMERO-DB to address this need. PRIMERO-DB has the same Health Insurance Portability and Accountability Act (HIPAA) securities as EMR data and is available from any web-enabled device, providing user-specific permissions to authorized users. Additionally, the COVID-19 pandemic coincided with the start of PRIMERO recruitment. With pandemic control measures in place and OB/GYN offices limiting their indoor capacity, recruiters lost the advantage of building the rapport provided through face-to-face interactions. As recruitment continued throughout the evolving pandemic, the generalizability of our findings to babies born before and after the pandemic will need to be evaluated.

Creating a birth cohort uniquely positions us to answer questions about the early-life origins of health and disease. We aim to leverage this rare opportunity to study the broader health of children over the next decades. Given adequate support, long-term prospective follow-up of PRIMERO participants will enable studies of numerous exposures and outcomes and enable the identification of biomarkers that may predict outcomes well before their traditional diagnosis. This information collectively promises insights into potential pathways that may be targetable in efforts to prevent illnesses and the subsequent development of childhood diseases such as asthma.

## Data Availability

Access to data and samples generated within the PRIMERO study is governed by a set of guidelines aimed at fostering collaboration and ensuring the integrity of the research endeavor. Data access is evaluated on a case-by-case basis and is limited to PRIMERO-approved investigators. Approval as a PRIMERO investigator involves evaluation by all three Principal Investigators (PIs; Drs. Sheppard, Seibold, and Rodriguez Santana) and recognition by the National Heart, Lung, and Blood Institute (NHLBI). External investigators interested in contributing to the mission, infrastructure, and sustainability of PRIMERO are encouraged to apply for investigator status.

## ACKNOWLEDGMENTS

The authors acknowledge the families for their participation and thank the numerous data collectors, recruiters, technicians, and hospital administrators for their support and participation in PRIMERO. The authors would also like to thank Iliany Cubero, Peter Deford, George Halley, Michael Montgomery, James Nolin, Bhavika Patel, Elizabeth Plender, Lisa Butler, and Narjis Kazmi. We also extend a special thank you to the San Francisco Chinatown Merchants Association for their generous donations of personal protective equipment during the COVID-19 pandemic.

## Abbreviations

API: asthma predictive index
CNP: Centro de Neumología Pediátrica
COVID-19: coronavirus disease 2019
EMR: electronic medical record
EPR-3: Expert Panel Report 3
GxE: gene-environment
HIMA: Hospital Interamericano de Medicina Avanzada
HIPAA: Health Insurance Portability and Accountability Act
HRV: human rhinovirus
IIAQ: Initial Illness Assessment Questionnaire IRB Institutional Review Board
ITFQ: Illness Tracking and Follow-up Questionnaire
LRI: lower respiratory tract illnesses
mAPI: modified asthma predictive index
MDIQ: Medical Doctor Illness Questionnaire
NAEPP: National Asthma Education and Prevention Program
NIH: National Institutes of Health
NJH: National Jewish Health
non-RI: non-respiratory tract illness
OB/GYN: obstetric/gynecologic
OSMB: Observational Study Monitoring Board
PRAM: Pediatric Respiratory Assessment Measure
PRIMERO: Puerto Rican Infant Metagenomic and Epidemiologic Study of Respiratory Outcomes
qPCR: quantitative polymerase chain reaction
REDCap: Research Electronic Data Capture
RI: respiratory illness
RNA: ribonucleic acid
RSS: Respiratory Severity Score
RSV: respiratory syncytial virus
SMS: short message service
UCSF: University of California, San Francisco
URI: upper respiratory tract illness

