## Supplemental Text and Figures for "The Puerto Rican Infant Metagenomic and Epidemiologic Study of Respiratory Outcomes (PRIMERO): Design and Baseline Characteristics for a Birth Cohort Study of Early-life Viral Respiratory Illnesses and Airway Dysfunction in Puerto Rican Children"

**SUPPLEMENTARY TEXT**

**Supplemental Text 1: Data Collection and Management**

Given the scale and complexity of the study (e.g., coordinating recruitment visits, study visits, and the unique procedures associated with each visit), the team developed an electronic record-keeping software *de novo* to track these tasks for all participants as they progress through each stage of the study. The system, PRIMERO-DB, is used to manage recruitment, surveillance, and follow-up activities, and is implemented with the same Health Insurance Portability and Accountability Act (HIPAA) securities as electronic medical record (EMR) data. Potential participants were first entered into PRIMERO-DB when recruiters informed pregnant women about the study. Women interested in the study were asked to offer their name, phone number, and expected due date (stage 1 consent must have occurred prior to the child’s birth). This information was entered into a tracking log in PRIMERO-DB, which in turn helped recruiters to prioritize informed consent and eligibility determination on a daily basis by providing a list of women sorted by due date. Upon presentation of the mothers to labor and delivery, PRIMERO-DB was used to identify consented mothers and manage the flow of tasks. Admitted women were cross-referenced against the PRIMERO-DB tracking log to identify consented and eligible mothers. Once the child was born, PRIMERO staff recorded the collection of cord blood from consented participants by scanning barcoded collection tubes into PRIMERO-DB. This in turn notified laboratory technicians to prepare for the arrival of samples and to scan them as received once they arrived at the laboratory. Reaffirmation of consent, determination of infant eligibility, and administration of the baseline questionnaire were also conducted through PRIMERO-DB. Automated weekly text/email messages asking participants about their child’s respiratory health, and their responses (or lack of) are sent through, and automatically recorded in PRIMERO-DB once the mother and child leave the hospital and enter the illness surveillance phase of the PRIMERO study.

The UCSF Research Electronic Data Capture (REDCap) system is used to collect, transfer, store, and manage recruitment and annual follow-up visit data. REDCap is a professionally managed, secure, HIPAA-compliant, web-based system for building and managing web-based research.

The UCSF Data Coordinating Center (DCC) monitors study protocol compliance and ensures data quality through range and logic checks at time of data entry, and through quality control assessments of the study database on a regular basis. PRIMERO staff are assessed for data completeness, data entry error rates, and internal consistency of collected data. Staff receive regular feedback on these metrics and opportunities for continued training on study protocols as necessary.

**Supplemental Text 2: Core Facilities and Sample Handling**

PRIMERO activities are coordinated through three core facilities:

Recruitment and Follow-up Core

CNP is the recruitment and follow-up core and coordinates activities between recruiters, data collectors, and staff that perform follow-up activities. CNP houses the text messaging and call center for RI surveillance and study visit scheduling. For PRIMERO, CNP constructed a negative-pressure biosafety level 2 laboratory with the capabilities of 1: processing, culturing, and storing nasal swab specimens, and 2: processing and isolating buffy coat and plasma samples for cryopreservation from maternal and infant cord and peripheral blood samples. Additionally, as a result of the COVID-19 pandemic, the CNP constructed a negative-pressure clinical examination room for patient visits.

Data Coordination Core

The UCSF Asthma Collaboratory is the data coordinating center (DCC) and the IRB of Record for all sites. The DCC helps collect, monitor, integrate, and distribute information for PRIMERO. The DCC generates and distributes data collection forms and protocols and maintains a coded data set on UCSF servers.

UCSF Laboratory Core

Biological specimens from the child are obtained from cord blood (cord blood mononuclear cells [CBMCs], plasma, deoxyribonucleic acid [DNA], and ribonucleic acid [RNA]) and during the year 2 annual visit (serum, plasma, DNA, and RNA); biological specimens from the mother (plasma, DNA, and RNA) are collected 24 hours postpartum. CBMCs and plasma extractions are performed at CNP. All biological specimens are coded and shipped to the UCSF Pediatric Asthma Specimen Bank (IRB# 10-00085) on dry ice. UCSF utilizes a Microsoft Access database to manage all biological specimens. Blood tubes are further processed at UCSF for DNA utilizing the Wizard® Genomic DNA Purification Kit (Promega, Fitchburg, WI).

NJH Airway Biobank Core

Two coded nasal airway swab samples are collected from each visit at CNP for 1: nucleic acid isolation, and 2: cryopreservation. Following collection in appropriate buffers optimized for downstream analyses, airway specimens are shipped to the NJH Airway Biobank Core in Denver, Colorado. For nucleic acid isolations, NJH uses a modified DNA/RNA isolation protocol using a Beckman Coulter Automated i7 Liquid Handling Workstation fitted with the Data Acquisition and Reporting Tool (DART) software for sample tracking and extraction of both DNA and RNA biomolecules prior to banking. Swabs collected for cryopreservation are banked in liquid nitrogen for downstream culture, *in vitro* experiments, and single-cell RNA sequencing (scRNA-seq) outcomes and analyses. CNP and NJH utilize both FreezerWorks and LabGuru electronic laboratory management systems to maintain sample tracking of shipments between sites and for extraction and inventory management of all PRIMERO biospecimens.

Addressing Operational Challenges for a Remote Study Site

For investigators based on the U.S. mainland, operating a study site removed from the continental U.S. can amplify the challenges of conducting a research study. We leveraged our experience working in Puerto Rico since the late 1990s to obviate some of these challenges. For example, blood samples from an individual are divided into two separate shipments and the second shipment is not sent until the first shipment has been received. In the event that a shipment is delayed while in transit and samples are lost because they were not maintained at the proper temperature (e.g., ice packs have fully melted), the second batch will not be sent until the reasons for the delay have been addressed. Past solutions have included switching couriers, adding more ice packs, and re-training staff. We also keep abreast of current events, including civil disturbances and weather phenomena. We have installed a -80°C freezer and self-sustaining liquid nitrogen storage capabilities at CNP to delay shipment of study samples if we anticipate that significant events may have the potential to affect a proper delivery. Laboratory information management system software is used to track the location and status of all study samples in real time.

Procurement Strategies

Procurement of study materials (nasal swabs, blood tubes, personal protective equipment, etc.), had been challenging with pandemic-related interruptions to supply chains. Thus, we have been diligent in securing new and back-up vendors, and in the stockpiling of enough supplies to allow study operations for the PRIMERO study to continue for at least two months.

**Supplemental Text 3: Respiratory Illness Surveillance**

Initial Illness Assessment

Mothers who report signs of illness in their child are contacted by project staff via phone. Project staff administer an Initial Illness Assessment Questionnaire (IIAQ), a high-sensitivity, low-specificity screening questionnaire designed to capture all LRIs based on the presence of symptoms and/or having seen a healthcare provider because of the illness. Symptoms suggestive of LRIs, URIs, and non-respiratory tract illnesses are described briefly in Supplemental Text 3. All likely LRIs are referred for an in-person clinic visit. The PRIMERO study protocol planned for one in-clinic URI assessment per child per year. URI visits were briefly paused under Puerto Rico’s COVID-19 stay-at-home mandate from March 15, 2020 to May 3, 2020, resuming after mandates were lifted.

Respiratory Illness Visits

During in-person visits, a clinical evaluation including a Medical Doctor Illness Questionnaire (MDIQ) and nasal swab are completed. The Pediatric Respiratory Assessment Measure (PRAM) and the Respiratory Severity Score (RSS) are administered to obtain a clinically standardized measure of the child’s illness severity. A final determination of URI versus LRI is recorded on the MDIQ based on the physician’s assessment, PRAM, and RSS scores.

Illness Follow Up

All RIs are tracked using the Illness Tracking and Follow-up Questionnaire (ITFQ) administered by phone following a RI visit or IIAQ without a scheduled face-to-face visit. PRIMERO staff conduct weekly ITFQs to document the trajectory of the RI and to help determine the current state of the illness, presence and severity of symptoms, and occurrence of various illness-related clinical events (e.g., diagnosis of bronchiolitis; prescription of oral steroids; illness-related hospitalizations). Once a RI is determined to have resolved, the child is returned to regular weekly SMS-text/email surveillance messaging.

Determination of Illness Type and Severity

An illness is classified as an LRI based on specific criteria, including the presence of clinician-assessed wheeze, accessory muscle use, oxygen saturation below 92%, PRAM score exceeding 3, RSS of 4 or higher, or self-reported symptoms consistent with an LRI, such as wheezing, cough interfering with daily activities, cough disrupting sleep, fast breathing, or gasping for air. LRIs are dichotomized as mild/moderate or severe. LRIs are classified as severe if the illness requires hospitalization or prescription of oral steroids, or if the illness PRAM score is in the severe range (8 to 12). Symptoms suggestive of a LRI include a cough that interferes with daily activities, the presence of wheezing/whistling in the chest, fast breathing or gasping for air, and sleep disturbed by cough, wheeze, or difficulty breathing. Participants reporting only to have a runny/plugged nose, sneezing, and/or a mild cough are considered most likely to have an upper respiratory tract illness (URI). Participants not reporting any respiratory symptoms are considered to have a non-respiratory tract illness (non-RI) and are advised to continue to monitor their child and seek care with a healthcare provider should their child’s illness worsen.

**Supplemental Text 4: Secondary Respiratory Illness outcomes and Research Questions**

Since we will have viral qPCR results from all documented LRIs for common respiratory virus species (HRV-A, HRV-B, HRV-C, RSV, metapneumovirus, parainfluenza, influenza), and prior results suggesting that the species of virus responsible for the infection may drive asthma risk, we will examine the following virus-informed secondary outcomes:

1. Early-life viral RI outcome. We will use viral qPCR results from all documented LRIs for common respiratory virus species (HRV-A, HRV-B, HRV-C, RSV, metapneumovirus, parainfluenza, influenza) to reclassify the early-life RI outcome by confirmed viral LRIs. Specifically, new groups will be defined as follows: Group 1: no viral LRIs; Group 2: at least one mild-moderate viral LRI but no severe viral LRIs; Group 3: at least one severe viral LRI.
2. Early-life HRV or RSV RI outcome. The relationship between early-life viral LRIs and asthma is strongest for RSV and HRV. Therefore, we will reclassify the early-life RI outcome as follows: Group 1: no HRV or RSV LRIs; Group 2: at least one mild-moderate HRV or RSV LRI but no severe HRV or RSV LRIs; Group 3: at least one severe HRV or RSV LRI.
3. Early-life HRV RI outcome. To identify specific effects of HRV, we will reclassify the early-life RI outcomes as follows: Group 1: no HRV LRIs; Group 2: at least one mild-moderate HRV LRI but no severe HRV LRIs; Group 3: at least one severe HRV LRI.
4. Early-life RSV RI outcome. To identify specific effects of RSV, we will reclassify the early-life RI outcomes as follows: Group 1: no RSV LRIs; Group 2: at least one mild-moderate RSV LRI but no severe RSV LRIs; Group 3: at least one severe RSV LRI.

We will perform six secondary analyses designed to answer the following research questions involving the 4 alternate, viral-informed reclassifications of the early-life RI outcomes.

A. What are the genetic determinants of airway gene expression at birth, in illness, and at two years of age?
B. What is the prospective association of newborn airway gene expression with early-life RI outcomes?
C. What is the association of year two airway gene expression with early-life RI outcomes?
D. What changes in airway gene expression from birth to two years of age are a consequence of early-life RI outcomes?
E. What is the association of early-life RI outcomes with asymptomatic virus infection at two years of age?
F. What is the association of early-life RI outcomes with the modified asthma predictive index (mAPI) outcome at two years of age?

**Supplemental Text 5: COVID-19 Precautions (Risk Mitigation Plan)**

The risk mitigation plan initiated enhanced safety measures for PRIMERO staff when interacting with participants and limited face-to-face interactions to medically necessary visits. Specifically, study activities related to birth and LRI visits were allowed to proceed since participants would already be present in a clinical setting for these medical events. Pre-delivery consenting was permanently switched to virtual (electronic signed consent is obtained via DocuSign®), and visits for URIs were suspended from March 15, 2020 to May 3, 2020.

Consenting

Due to the COVID-19 pandemic, we amended our consenting process to include an electronic component. We moved from a hardcopy PRIMERO Information Packet to an electronic version. Partnered obstetricians were also provided with an electronic copy of the PRIMERO Information Packet to help with distribution to potential participants. Stage 1 consent was modified to obviate exposure risk by providing potential participants with HIPPA-compliant electronic consent forms via DocuSign®, which they were instructed to sign and return electronically. During the signing process, recruiters were on the phone with the potential participant to field any questions. Stage 2 reaffirmation (postnatal) was administered at the bedside by a recruiter in full personal protective equipment (PPE). Despite these precautions, an in-person meeting presented an exposure risk to participants and research staff. As such, the recruitment site hospital (HIMA) implemented universal screening for SARS-CoV-2 among pregnant mothers scheduled to deliver at HIMA.

Screening of mothers and babies at HIMA

As per hospital procedure, all mothers scheduled to deliver at HIMA were screened for SARS-CoV-2 antibodies using a rapid test. If the screening was positive, a PCR-based test was performed to determine if the patient had an active infection. If the mother had an active infection, all contact with her was conducted through hospital personnel assigned to the mother’s care. The protocol at HIMA for PCR-positive women presenting for labor was to isolate them in a negative pressure room within a dedicated COVID-19 ward that was on a different floor from labor and delivery. Babies born to PCR-positive mothers were also screened for SARS-CoV-2 by PCR. Treatment of mothers and children for COVID-19 took priority over PRIMERO research procedures, and continuation of birth procedures was considered on a case-by-case basis so as not to interfere with clinical care.

Birth procedures

Collection of maternal peripheral blood and administration of the baseline questionnaire was conducted at the bedside by staff. No maternal blood was collected if a mother was PCR-positive for SARS-CoV-2. If a newborn was anticipated to be infected with SARS-CoV-2, the child’s nasal swab was obtained immediately after birth. Results from PCR testing were available within 24 hours. If the swab was negative, PRIMERO staff proceeded as per study protocol and obtained nasal swabs. Newborn nasal swabs were obtained in a separate room by the research staff (nurse, medical technologist, or physician). No nasal swabs were obtained from babies with positive PCR results. PRIMERO staff arranged an agreement with HIMA whereby hospital staff transport the placenta from the operating room and labor suite to a separate room dedicated for PRIMERO staff to perform cord blood extractions. This minimized exposure between PRIMERO staff, participants, and hospital employees. Cord blood collection continued on all participants. Staff wore full PPE as required by HIMA and CNP guidelines and practiced appropriate social distancing when possible.

Illness surveillance visits

Clinic visits for RIs present an exposure risk to participants and research staff. Study visits that were not medically necessary (upper respiratory tract illnesses in the first two years of life) were deferred during the COVID-19 pandemic from March 2020 until May 2020. Visits for URIs resumed in June 2020. Clinic visits for LRIs continued to occur regardless of study protocol because they are considered medically necessary. Research activities were performed as per protocol after clinical care had been administered for all visits that are medically necessary, with appropriate COVID-19 precautions for staff and participants as described in Supplementary Table 1. If a participant is suspected of having or confirmed to have SARS-CoV-2, a separate protocol is used (“Persons suspected or confirmed infected with SARS-CoV-2,” below).

Annual follow-up visits

Although it is uncertain how much the COVID-19 pandemic affected annual visits, study staff followed appropriate COVID-19 protocols in accordance with the local government and hospital regulations when conducting these visits.

Persons suspected or confirmed infected with SARS-CoV-2

PRIMERO staff avoided face-to-face interactions and sample collections from potential or current participants who were suspected or confirmed to be infected with SARS-CoV-2 (by PCR testing), regardless of whether they were symptomatic. Face-to-face interactions resumed for individuals with PCR-positive results after they have had a negative PCR test for SARS-CoV-2. As the pandemic evolves and children age, it is possible that a child in PRIMERO will become infected with SARS-CoV-2. If a face-to-face encounter is required and COVID-19 is suspected, the child is referred for PCR testing and must have a negative result before being seen at CNP (SARS-CoV-2 testing is free in Puerto Rico). Similar practices are being followed for children in PRIMERO: defer medically unnecessary visits; use PPE for all face-to-face encounters; if infection is suspected, refer for PCR testing and medical care; avoid sample collection and face-to-face encounters while children are PCR-positive; collect illness characteristics via electronically administered illness questionnaires; resume face-to-face encounters for medically necessary visits after a negative PCR result.

**SUPPLEMENTARY FIGURES**

**Supplemental Figure 1.** Example SMS text received (left panel) and linked page (right panel) by which PRIMERO participants respond to weekly message enquiring about their child’s health.

| 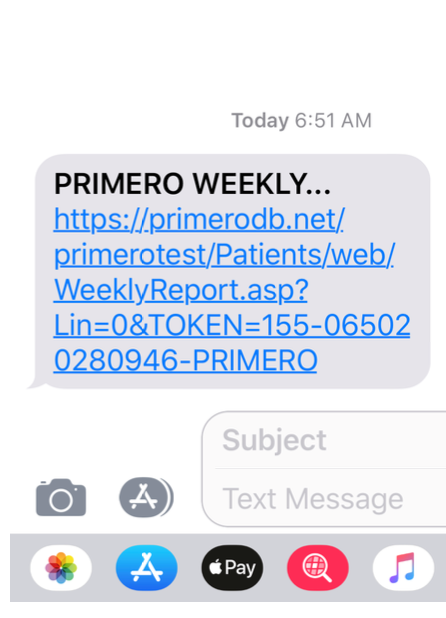 | 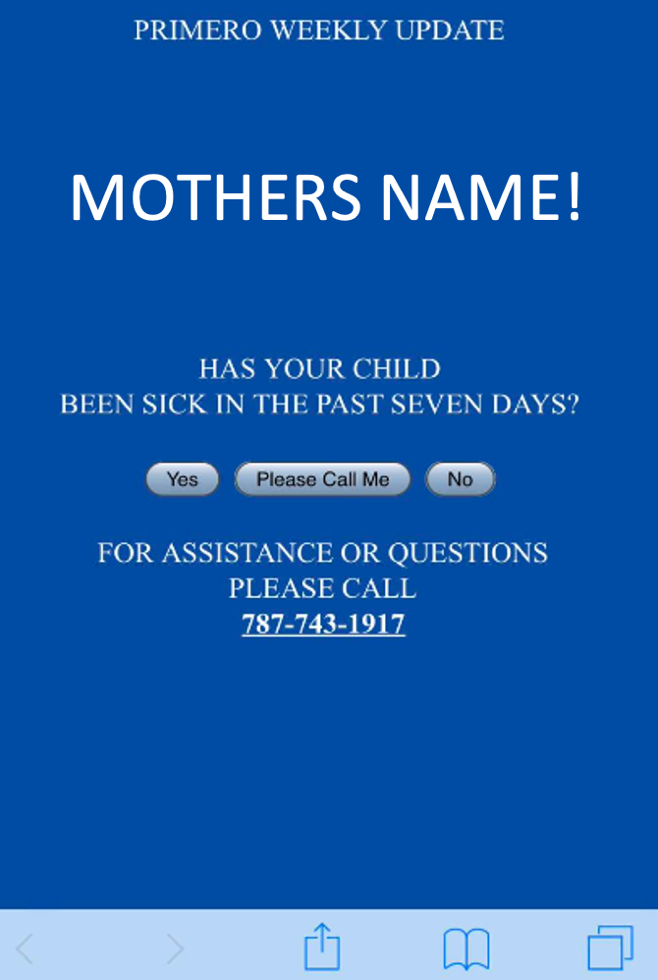 |
| --- | --- |

**Supplemental Figure 2.** Example participant engagement through study website updates, and personalized greetings. All engagement materials are available and distributed in the participant’s preferred language.

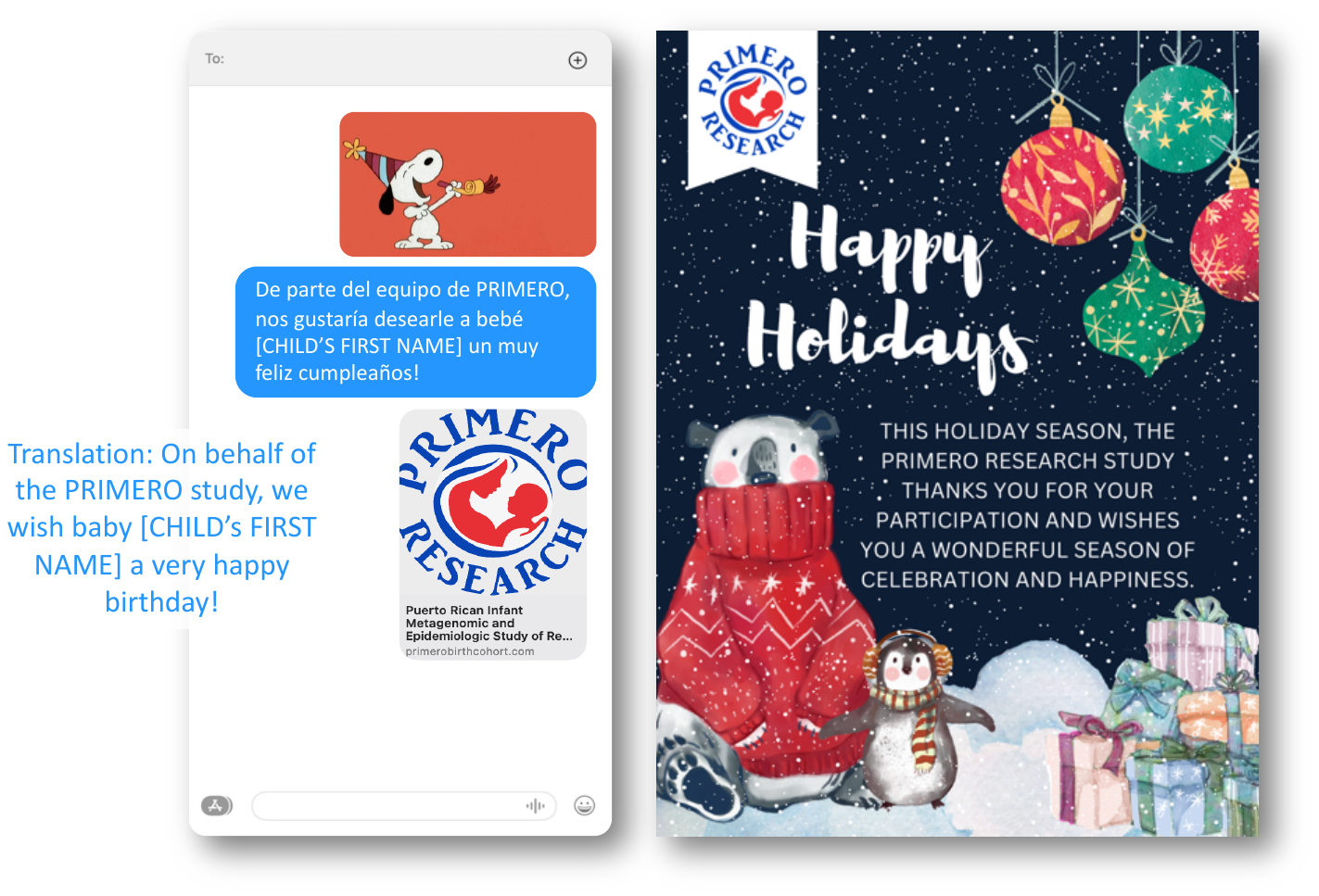

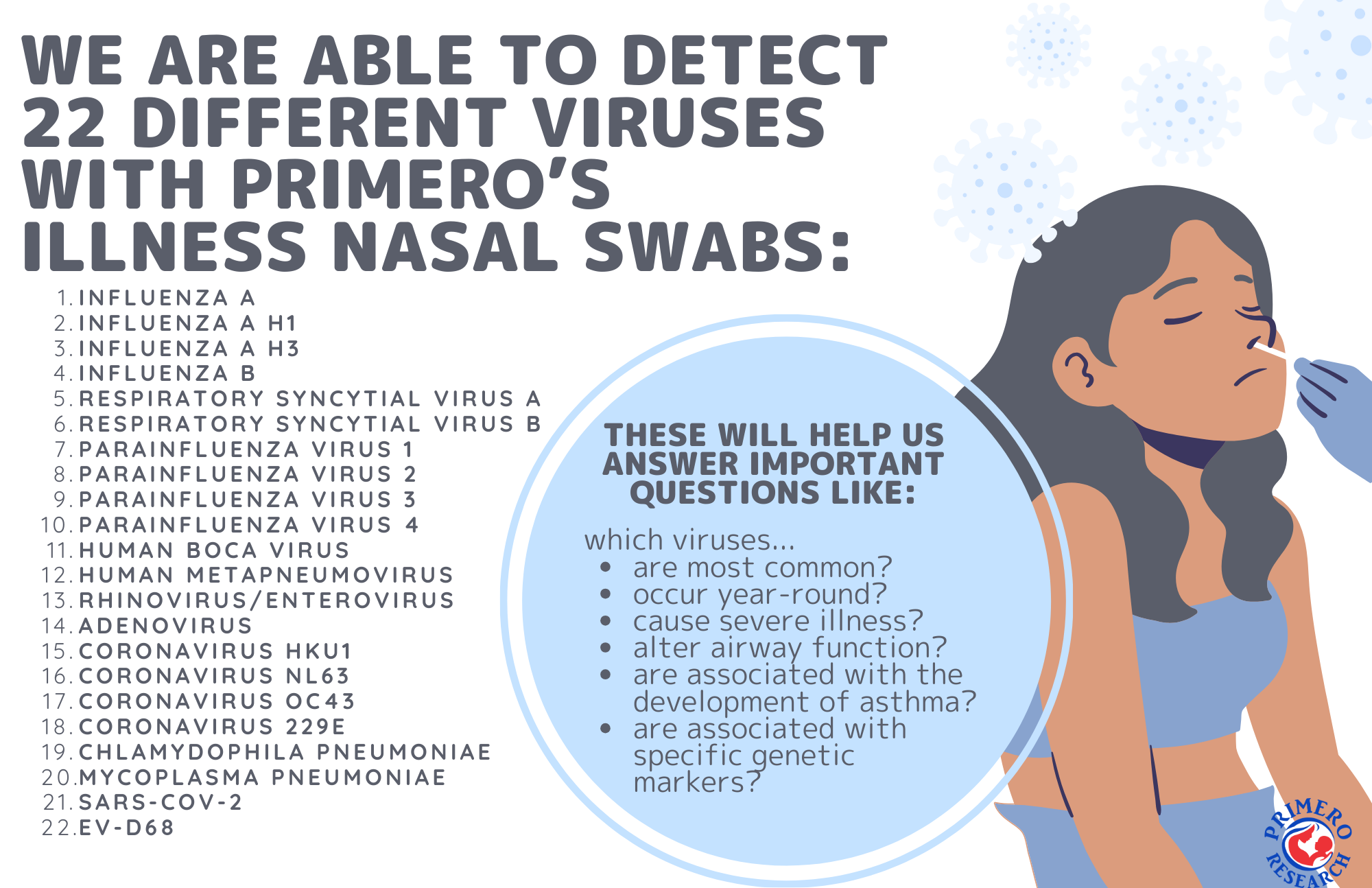

**Supplemental Figure 3.** Monthly and cumulative PRIMERO enrollment from March 2020 to June 2023

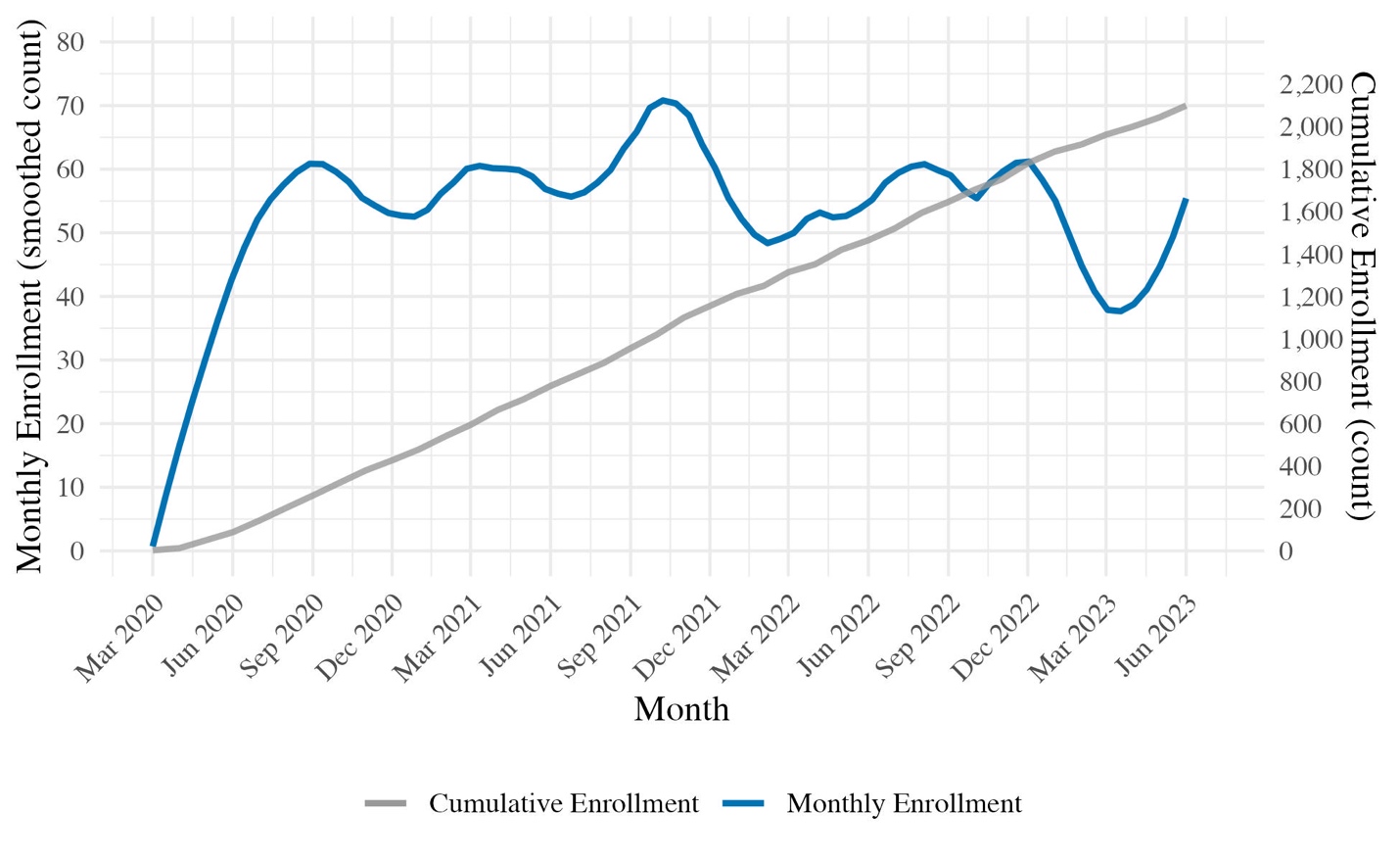

The inaugural eligible infant entered the study in March 2020, and a median of 55 infants were enrolled each month thereafter. The PRIMERO cohort consists of 2,100 term healthy infants, with enrollment concluding in June 2023. Smoothed count: the Locally Weighted Scatterplot Smoothing (LOESS) method was used to fit a smooth curve through the monthly enrollment count data points.
